# Surface sampling for mpox virus in multiple healthcare settings in Sierra Leone, June 2025

**DOI:** 10.1101/2025.09.16.25335757

**Authors:** Michaella Jaba, Emmanuel J. Siadu, Emmanuel S. Kamanda, Christina Frederick, Laurens Liesenborghs, Megan Halbrook, Nicole A. Hoff, Placide Mbala-Kingebeni, Isaac I. Bogoch, Souradet Y. Shaw, Anne W. Rimoin, Jia B. Kangbai, Jason Kindrachuk

## Abstract

**Background:** As mpox virus (MPXV) has continued to expand geographically, critical knowledge gaps in environmental persistence in resource-limited healthcare settings remain. Despite evidence of fomite-mediated transmission, no empirical data exist on surface contamination in West African hospitals during active mpox outbreaks.

**Methods:** In this cross-sectional study, we conducted a systematic environmental surveillance study at two major hospitals in Sierra Leone (Connaught Hospital, Freetown; Bo Government Hospital, Bo) during peak transmission (June 2025). A total of 89 high-contact surfaces were sampled across clinical and non-clinical zones using standardized protocols. MPXV DNA was extracted via robotic MagMax protocols and detected through quantitative real-time PCR targeting the B6R gene.

**Results:** Overall, 13.5% (12/89) of surfaces tested positive by PCR for MPXV, with geographic variation: Freetown (14.0%, 7/50) vs. Bo (12.8%, 5/39). Cycle threshold values (32.34–39.86) indicated low-to-moderate viral genomes. Critical contamination hotspots were identified, with doors representing 42% (5/12) of positive samples; predominantly ward entrances, staff offices, and bathrooms. Patient beds and clinical instruments constituted secondary risk zones (8.3% each).

**Conclusions:** This first-in-region study demonstrates quantifiable MPXV genomes from surfaces in Sierra Leonean healthcare facilities, providing support for patients with mpox, highlighting areas for infection prevention and control (IPC) considerations. The predominance of doors as high-risk fomites underscores the need for targeted disinfection protocols. Our findings establish environmental surveillance as a vital component of mpox control in clinical settings and provide evidence for IPC resource prioritization, including enhanced disinfection of high-touch surfaces and integration of IPC monitoring into national outbreak response frameworks.

## INTRODUCTION

Mpox, formerly known as monkeypox, is an emerging zoonotic viral disease caused by mpox virus (MPXV) [1] in the *Orthopoxvirus* genus of the *Poxviridae* family. MPXV is divided into two phylogenetically distinct clades: Clade I (endemic to regions of Central Africa) and Clade II (endemic to regions of West Africa), with both clades further subdivided into subclades a and b [2]. Historically endemic to Central and West Africa, mpox gained renewed attention following the global outbreak of Clade IIb in 2022 [3]. This outbreak was typified by sustained human transmission and concentration of infections among dense sexual networks and included substantial changes in clinical and epidemiological characteristics [4,5]. The rapid increase in cases and spread of Clade IIb MPXV led to the first public health emergency of international concern (PHEIC) declared for mpox by the World Health Organization (WHO) in July 2022 [6].

Healthcare facilities act as a sanctuary for medical treatment, yet a potential source for pathogen propagation. A good example is the 2014 outbreak of the Ebola Virus Disease (EVD) in Sierra Leone, amongst other viral hemorrhagic fever outbreaks, which led to gaps in infection prevention and control (IPC) standards across many public health facilities [7]. Post-Ebola funding for IPC infrastructure showed significant improvement, but environmental hygiene and waste management processes and surface disinfection, along with clinical area maintenance, have shown a decrease in support for sustained operation of these activities [8]. Investigations of Clade IIb MPXV showed evidence for altered clinical and epidemiological characteristics, including within clinical settings, suggesting contaminated healthcare materials could facilitate indirect transmission pathways [9,10]. Thus, fomites could facilitate transmission opportunities, especially in health settings where resource limitations could negatively impact surface disinfection procedures [9,10].

Evidence from recent mpox outbreaks in high-income countries shows that MPXV DNA can persist on surfaces such as bed linens, doorknobs, and medical instruments for extended periods, even after a patient is discharged [11]. Recent data from a UK specialist hospital revealed that MPXV DNA was detected on two-thirds of sampled surfaces. At the same time, several surfaces also showed the presence of a virus that researchers identified as viable [12]. Morgan et al. discovered that MPXV persisted in a patient’s residential area for 15 days following symptom disappearance [10]. The aggregation of these discoveries underscores the essential role played by healthcare environmental contamination assessment as front-line workers face inconsistent IPC implementation, particularly in resource-limited settings [13].

Currently, there are critical knowledge gaps regarding MPXV surface contamination in healthcare settings within regions of Africa affected by mpox public health emergencies, including Sierra Leone. This includes the lack of first-hand hospital surface swab data for MPXV detection. Furthermore, healthcare managers need to comprehend surface-driven disease spread possibilities for better controlling infection prevention guidelines and for correctly distributing cleaning materials, as well as safeguarding healthcare workers, staff, patients, and visitors. The collection of comprehensive surface samples at medical institutions provides operational data on contamination vulnerabilities, alongside the discovery of important locations of pathogen contamination [14].

This investigation aimed to address the existing gaps for MPXV surface contamination in healthcare settings in Sierra Leone at two geographically distinct study sites: Connaught Hospital (Freetown) and Bo Government Hospital (Bo), in June 2025. We used standardized molecular screening methods to establish hospital surface MPXV positivity rates, determine risk areas inside clinical spaces, and generate adaptable infection prevention and control strategy recommendations.

## METHODS

### Study Setting and Location

The study was conducted in Freetown and Bo, urban epicenters representing the Western Area Urban District and Bo District, respectively. In this study, we selected healthcare environments, specifically high-contact surfaces within several medical wards and departments in two major urban public hospitals in Sierra Leone: Connaught Hospital in Freetown and Bo Government Hospital in Bo. These locations were strategically chosen due to their health infrastructure diversity and ongoing mpox surveillance operations supported by the Ministry of Health and Sanitation. Both hospitals serve as regional referral centers, offering a relevant setting for exploring environmental persistence of the mpox virus within low-resource healthcare environments that reflect broader regional IPC capacities [15].

### MPXV DNA extraction PCR testing

A robotic MagMax DNA Multi-Sample Ultra Kit was used to extract and purify DNA from all 89 surface swab samples. The swabs were aliquoted into a 96-deep well plate. 200 μL of PK mix was added to each well that contained the swab. PK mix was made up of 8 μL of proteinase K and 192 μL of PK buffer. The plate was sealed with a clear adhesive film and was shaken for 5 minutes at a speed of 950 rpm. After which, it was incubated for 20 minutes at a temperature of 65 °C. During incubation, 3 sets of washes were prepared according to the manufacturer’s manual, followed strictly for subsequent steps and all procedures [16]. For qPCR, the MPXV B6R gene was targeted. The real-time PCR thermal cycler (Bio-Rad CFX96) was programmed for a run of 20 μL per sample, for a total amplification time of 60 minutes. After the run, the data were transferred into a pen drive and imported into a computer for analysis [17, 18].

### Analytical Methods

Descriptive statistics (e.g., frequency of positive samples) were used to summarize contamination patterns. Data were stratified by location, surface type, and Ct value (with Ct <35 used to infer higher contamination loads). No imputation or correction for missing data was required. Results were visualized using bar graphs and heatmaps to facilitate pattern recognition across surface categories [19].

No alternative interventions were trialed. However, various hospital environments were compared to evaluate contamination differences. Comparators included surface types (e.g., metallic vs. plastic), functional areas (clinical vs. administrative), and locations (Bo vs. Freetown). This comparative stratification provided indirect evaluation of IPC effectiveness in different spatial and procedural contexts [22].

## RESULTS

This investigation adopted a health system perspective, focusing on operational-level IPC gaps. From this perspective, the study aimed to evaluate environmental contamination as a cost-invisible contributor to mpox transmission risk. While no direct monetary costing was applied, the findings will directly inform decisions around resource allocation for IPC protocols, especially disinfection strategies and protective measures for healthcare workers. This evaluation was based on a single observational study involving primary field sample collection and laboratory-based molecular diagnostics. A total of 89 surface swab samples were randomly collected in two major districts in Sierra Leone: Bo, where 39 surface samples were collected between 02 June 2025 and 15 June 2025, and in Freetown, where 50 surface samples were collected between 12 June 2025 and 25 June 2025. In Bo, samples were collected at the Bo government hospital from various hospital surfaces, including bathrooms, doorknobs, patient beds, corridors, wards, and offices, using a sterile polyester-tipped applicator swab, as well as at the Connaught hospital in Freetown. This approach provided sufficient resolution to detect sub-threshold viral DNA contamination across different hospitals. The primary outcome was binary: presence or absence of MPXV DNA on surfaces, determined via real-time PCR. Whilst this does not directly measure disease transmission or clinical impact, it serves as a vital representation for environmental risk and potential fomite-mediated exposure. This is particularly relevant in settings with vulnerable patient populations and inconsistent IPC adherence [20].

A total of 89 surface swabs were collected and tested using a validated qPCR protocol using the B6R gene for virus detection: an established diagnostic marker for MPXV [21]. All sampling surfaces were swabbed completely, and the samples were transported using universal transport media. Surface swabs were obtained from both clinical (e.g., delivery beds, medical instruments) and non-clinical (e.g., door handles, bathrooms) areas. This stratification allowed for subgroup analysis comparing contamination in patient care versus auxiliary spaces, helping assess where mpox viral DNA is most likely to persist. The following assumptions were made: detection of MPXV DNA via qPCR indicated potential environmental contamination, sample locations were representative of typical hospital use patterns during the surveillance period, and negative surface results do not rule out earlier contamination or intermittent cleaning.

Out of the 89 surface swab samples collected from healthcare environments in Bo and Freetown, 12 tested positive for mpox virus DNA, representing a positivity rate of 13.5%, while 77 (86.5%) tested negative (Figure 1). Positive Ct values ranged between 32.34 and 39.86 (Table 1), suggesting moderate viral DNA load on those surfaces. Although Ct values are not a direct proxy for infectivity, consistently lower Ct values (<35) correlate with a higher likelihood of viable virus presence [23]. Among the 89 samples screened, 17 (19.1%) yielded measurable Ct values. Of these, 12 samples (70.6%) had Ct values <40 and were considered positive, while five samples (29.4%) had Ct values >40, consistent with background amplification or degraded DNA (Table 1). Geographical comparison revealed slightly higher contamination in the Freetown sampling location (7/50 samples; 14%) as compared to the site in Bo (5/39 samples; 12.8%) (Figure 1; Table 2). Surface-level analysis (Table 2) indicated that 42% (5/12) of positive results came from doors, particularly ward entrances, staff offices, and toilets. This was followed by patient beds, table surfaces, bathroom handles, and clinical instruments, each accounting for 8.3% of positives. From this data, 42% of the places of collection with positive tests were doors. The other places, like the bathroom, bed, and table, among others, comprised 8% each of the total samples that were positive for MPXV.

**Table 1:**
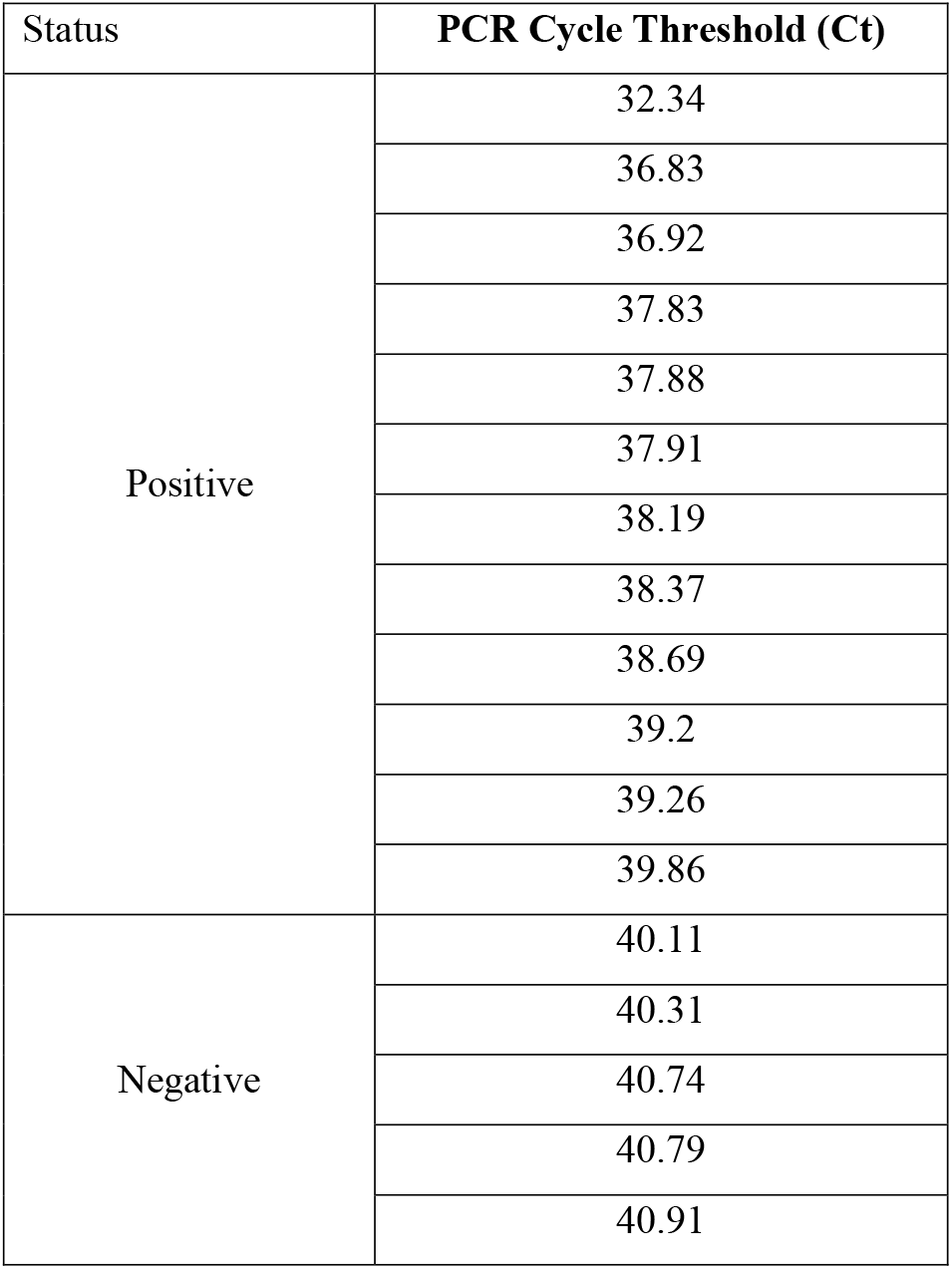
MPXV PCR-positive surface samples. Ct values for MPXV PCRs targeting the B6R gene.

| Status | PCR Cycle Threshold (Ct) |
| --- | --- |
| Positive | 32.34 |
|  | 36.83 |
|  | 36.92 |
|  | 37.83 |
|  | 37.88 |
|  | 37.91 |
|  | 38.19 |
|  | 38.37 |
|  | 38.69 |
|  | 39.2 |
|  | 39.26 |
|  | 39.86 |
| Negative | 40.11 |
|  | 40.31 |
|  | 40.74 |
|  | 40.79 |
|  | 40.91 |

**Table 2:** Identification of MPXV PCR positive surfaces and room locations.

| LOCATION | PLACE OF COLLECTION | PCR Result |
| --- | --- | --- |
| Bo | Lab office, bathroom | Pos |
|  | Female obstetrics and gynecology ward | Pos |
|  | Labor ward delivery bed & weighing scale | Pos |
|  | Eye clinic, vision lens and alcohol swab | Pos |
|  | Maternity theater, recovery bed and instrument trolley | Pos |
| Freetown | Female surgical ward ,main door | Pos |
|  | Male surgical ward, toilet door | Pos |
|  | Clinical office door | Pos |
|  | Sop entrance | Pos |
|  | Male medical ward, door | Pos |
|  | Male surgical ward, main door | Pos |
|  | Surveillance unit, table surface | Pos |

**Figure 1:**
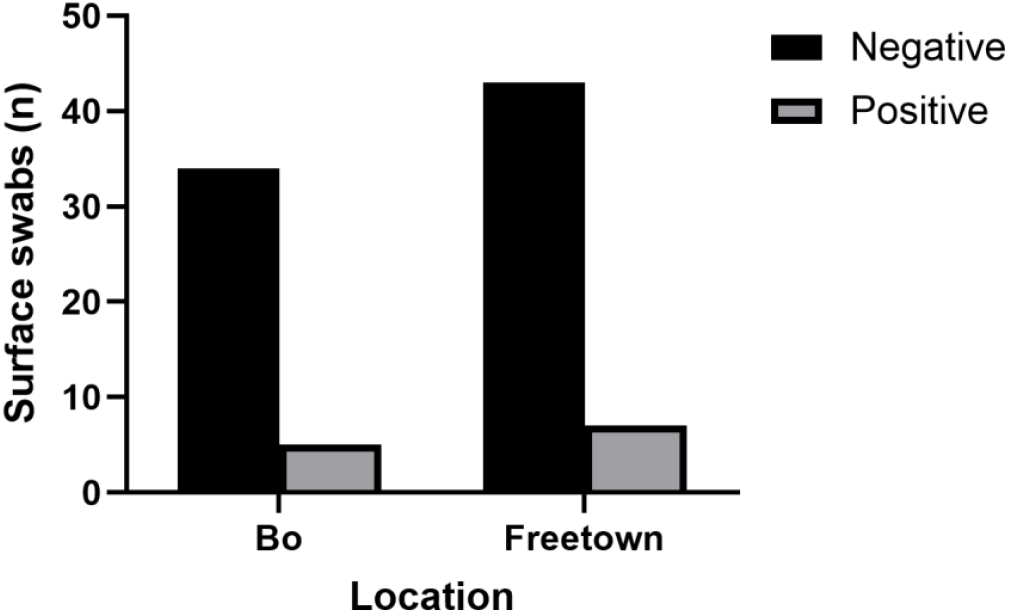
MPXV PCR results from surface sampling in Bo and Freetown, Sierra Leone. Total samples collected in Bo and Freetown in June 2025.

## DISCUSSION

This study represents the first systematic assessment of MPXV environmental contamination by surface swab and PCR analysis in healthcare facilities in Sierra Leone during the 2025 surveillance period. Using standardized surface sampling and molecular diagnostics, we identified a 13.5% overall positivity rate across high-contact surfaces in two major hospitals (Connaught Hospital, Freetown, and Bo Government Hospital, Bo). While the prevalence of surface contamination is lower than that reported in some high-income settings, the pattern and nature of contaminated sites suggest persistent fomite-related risks within low-resource healthcare environments.

The detection of MPXV DNA on 13.5% of sampled surfaces underscores the environmental persistence of the virus in active healthcare settings. Our findings are comparable to those from Gould et al., who reported a contamination rate of 19% in UK hospital isolation rooms during the 2022 outbreak [24]. Similar higher detection levels were observed in a Chinese study (52.66%) targeting isolation wards [25], while Pfeiffer et al. reported contamination rates exceeding 60% in mixed domestic and clinical environments [26]. The lower prevalence observed in our setting may partly reflect differences in sampling protocols, cleaning frequency, or environmental conditions such as humidity and temperature, which influence viral persistence. However, the presence of viral DNA on more than one in eight surfaces still represents a significant IPC concern, particularly in facilities where patient and staff movements are frequent, and cleaning practices are inconsistent.

The detection of MPXV DNA, even without confirmation of infectivity, aligns with global evidence indicating that orthopoxviruses are environmentally robust [27]. This reinforces the argument for routine environmental surveillance in African hospitals, which, despite limited resources, could act as an early warning system for nosocomial transmission risks.

Cycle threshold (Ct) analysis revealed that 70.6% of samples with measurable Ct values had Ct < 40, indicative of detectable viral DNA loads, with values ranging from 32.34 to 39.86. Prior studies suggest that Ct values below 35 are more likely to correlate with the presence of viable virus [23,28]. Morgan et al. documented MPXV DNA on surfaces for up to 15 days post-symptom resolution in residential settings, with similar Ct ranges [29]. Likewise, Atkinson et al. reported detection in domestic environments following imported cases into the UK, noting that some samples with lower Ct values yielded viable virus in culture [30].

While this study did not attempt viral isolation, the Ct profile observed here indicates that some contaminated surfaces could harbor infectious particles, particularly in high-contact zones. These results underline the potential for indirect transmission in healthcare facilities and the need for targeted disinfection of surfaces with repeated human contact. Importantly, Ct values should be interpreted cautiously; they quantify viral DNA, not infectivity, and can be influenced by swabbing technique, surface type, and time since contamination.

Geographical analysis showed slightly higher contamination rates in Freetown (14%) compared to Bo (12.8%). Although the difference is modest, it mirrors patterns from other infectious disease outbreaks where tertiary, high-throughput hospitals in urban centers exhibit greater environmental contamination [31]. This may reflect higher patient loads, greater diversity of clinical procedures, and more frequent contact between infected and uninfected individuals within confined spaces. Differences in IPC compliance, availability of cleaning materials, and staff training could also play a role [32].

A striking observation was that 42% of all positive samples were collected from doors, ward entrances, toilet doors, and office doors; followed by patient beds, bathroom handles, table surfaces, and clinical instruments. This distribution is consistent with Wojgani et al., who identified door handles as major contamination hotspots for multiple pathogens in hospital settings [33], and CDC guidelines, which classify them as high-risk fomites [34].

The prominence of doors as contamination points is unsurprising: they are touched by patients, visitors, and staff, often in rapid succession, and rarely disinfected between contacts. This is particularly problematic in wards without automated or hands-free entry systems. The other positive sites (delivery beds, recovery beds, and clinical equipment) also represent points of direct patient contact and should be prioritized for enhanced disinfection schedules. Installing hand hygiene stations near these surfaces, along with increased cleaning frequency, could significantly reduce environmental viral loads.

These results reinforce the persistence of viral DNA in healthcare environments and justify environmental sampling as a complementary surveillance tool, especially in LMICs where patient overcrowding, and IPC inconsistencies persist [13]. Furthermore, these findings suggest frequent recontamination due to repeated contact by patients, staff, and visitors, highlighting the need for hand hygiene stations and scheduled disinfection protocols at all doorways in clinical wards.

This research is not only scientifically significant, but it also contributes to global health security by supporting the development of early detection systems, informing infection control audits and hospital sanitation practices, and guiding national health policy formation. It supports global health security objectives through the development of early detection systems, infection control audits, and hospital sanitation, as well as national health policy formation. The global geographic expansion of mpox, along with rapid clinical and epidemiological changes, requires immediate programmatic upgrades for African healthcare facilities to strengthen environmental surveillance. The findings of this research can inform operational policies at the Ministry of Health and allied institutions, such as the National Public Health Agency (NPHA) in Sierra Leone, and guide resource prioritization for hospital disinfection and the training of healthcare workers.

Several limitations should be acknowledged. First, the study’s cross-sectional design provides only a temporal snapshot of environmental contamination, precluding assessment of persistence or the effects of cleaning interventions over time. Serial cross-sectional studies would provide more comprehensive data on viral survival dynamics in hospital environments. Second, the absence of viral culture means we cannot definitively determine whether the detected viral DNA represented an infectious virus. While prior literature suggests a correlation between lower Ct values and viability [23], confirmatory culture would strengthen conclusions about transmission risk. Third, the sample size, while adequate for exploratory purposes, was limited to two facilities, potentially restricting generalizability to other healthcare settings in Sierra Leone or West Africa. Finally, environmental conditions (e.g., temperature, humidity) were not systematically recorded, which may have influenced viral persistence and detectability. Despite these limitations, our findings are relevant to healthcare facilities in other low- and middle-income countries with similar structural and operational constraints. The identification of high-touch surfaces as key contamination sites aligns with IPC priorities worldwide and suggests that targeted interventions, such as more frequent disinfection of door handles and patient beds, could be both feasible and impactful in resource-limited environments.

## CONCLUSION

This study confirms environmental contamination by MPXV on high-touch hospital surfaces in Sierra Leone, with a 13.5% positivity rate. Doors emerged as the most contaminated surfaces, highlighting their role as potential fomites in healthcare-associated transmission. Moderate Ct values suggest the presence of viral DNA at levels warranting concern, even if not directly infectious. These findings stress the need for strengthened infection prevention protocols, especially routine surface disinfection and improved hand hygiene practices. Integrating environmental surveillance into national IPC strategies will be vital for controlling mpox and other emerging pathogens in resource-limited settings. This work also supports the integration of environmental surveillance into national mpox preparedness frameworks. Routine surface sampling could complement patient testing to detect early signs of nosocomial spread, triggering timely IPC interventions. In the broader context of global health security, such measures align with WHO’s emphasis on strengthening core public health capacities under the International Health Regulations [35]. Finally, our results reinforce the growing body of evidence that MPXV can persist in healthcare environments and that high-contact surfaces act as reservoirs for potential indirect transmission. The consistency of our findings with studies from Europe [24,25,30], North America [26,29], and Asia [36] suggests that environmental contamination is a universal feature of mpox outbreaks, albeit with variable prevalence depending on IPC infrastructure and outbreak stage.

## Data Availability

All data produced in the present work are contained in the manuscript

## AUTHOR CONTRIBUTIONS

JK, JBK, and AWR conceived the study and provided leadership support and guidance. MJ assisted with sample collection, designed the methodology, analyzed the results, and wrote the first draft. EJS and ESK received and recorded all samples. MJ and EJS processed all samples and performed all analyses. JBK coordinated the sample collection from the hospitals. All authors reviewed the manuscript.

## CONFLICT OF INTEREST

The authors declare no conflict of interest

## FUNDING STATEMENT

This work was funded by a Tier 2 Canada Research Chair in the Molecular Pathogenesis of Emerging and Re-Emerging Viruses for J.K., provided by the Canadian Institutes of Health Research (Grant no. 950-231498); and the International Mpox Research Consortium (IMReC), jointly funded by the Canadian Institutes of Health Research and International Development Research Centre (grant nos. MRR-184813).

## ETHICAL STATEMENT

The Sierra Leone Ethics and Scientific Review Committee (SLERC No. 010/05/2025) approved this study. Informed consent from mpox patients for confidentiality purposes was waived as collections and analyses focused on surface samples only.

## ACKNOWLEDGEMENT

The authors would like to acknowledge the staff at the Bo Government and Connaught hospitals, both in Freetown and Bo, for providing access for sample collection in various locations around the hospitals during these challenging times. The authors would also like to acknowledge the support provided by the Molecular Pathogenesis lab at the Department of Biological Sciences, Njala University, Njala Campus in Makonde, for all sample processing and analysis.

## Notes

### Competing Interest Statement

The authors have declared no competing interest.

